# Investigating the Determinants of Cervical Cancer Disease State Among Women Living with HIV at Cancer Disease Hospital in Zambia: A Retrospective Cross-Sectional Study

**DOI:** 10.64898/2026.08.10.26360062

**Authors:** Tembo Santos, Chileleko Mapiki, Maureen Mupeta Kombe, Michelo Charles

## Abstract

**Background:** Cervical cancer is the most common malignancy among Zambian women, and HIV co-infection accelerates disease progression. However, in the contemporary era of widespread antiretroviral therapy, the determinants of cervical cancer disease state among women living with HIV at Zambia’s primary oncology referral centre are not fully understood. This study aimed to investigate these determinants at the Cancer Disease Hospital in Lusaka.

**Methods:** An analytical cross-sectional study was conducted using medical records of 198 WLHIV with histologically confirmed invasive cervical cancer. Data on demographics, HIV status (CD4 count, viral load), and cancer characteristics (FIGO stage) were abstracted. Descriptive statistics, bivariate analyses, and multivariable logistic regression were performed.

**Results:** The prevalence of advanced-stage (Stage III/IV) cervical cancer was 34.4% (68/198), while 54.5% presented with Stage IIB disease. Metastatic disease was only 7.1% (14/198). The peak age of diagnosis was 40–49 years (51.5%). Median CD4 count was 484 cells/uL, and 89.4% had suppressed viral loads. No significant association was found between HIV disease status and advanced-stage cancer (CD4 <200: AOR 1.42, 95% CI 0.68–2.96; detectable viral load: AOR 1.38, 95% CI 0.71–2.68). Younger WLHIV aged 30–39 years had the highest proportion of advanced-stage disease (38.5%), as did peri-urban residents (40.0%) compared to Lusaka residents (30.0%).

**Conclusion:** In the ART era, HIV disease status is no longer the dominant determinant of cervical cancer stage among WLHIV at CDH who develop invasive cancer, likely due to effective immune reconstitution. However, persistent Stage IIB presentation indicates inadequate screening coverage. Younger WLHIV and peri-urban residents are at highest risk of late-stage diagnosis.

## BACKGROUND

Cervical cancer represents a critical and preventable global health challenge, with its burden disproportionately borne by women in low- and middle-income countries, particularly those in Sub-Saharan Africa (SSA) [9]. In Zambia, the disease is the most common cancer among women, presenting a significant public health crisis [10]. Zambia has the third highest burden of cervical cancer globally, with an incidence rate of 65.5 per 100,000 women and a mortality rate of 43.4 per 100,000 women, accounting for approximately 23% of all new cancer cases in the country [16,10]. The intersection of cervical cancer with the human immunodeficiency virus (HIV) epidemic creates a particularly severe synergistic health burden. Women living with HIV are six times as likely to develop cervical cancer compared to their HIV-negative counterparts [17,16].

HIV infection is a well-established risk factor for the development and aggressive progression of cervical cancer, primarily due to its role in compromising cell-mediated immunity, which is crucial for controlling persistent infection with high-risk human papillomavirus (hrHPV) [9]. A global meta-analysis demonstrated that women living with HIV have a pooled relative risk of 6.07 for cervical cancer, with the most affected regions being southern Africa and eastern Africa [9]. In southern Africa, an estimated 63.8% of women with cervical cancer are living with HIV, representing a substantial HIV-attributable cervical cancer burden [9].

The pathophysiological link between HIV and cervical cancer is primarily mediated through immunosuppression. HIV-induced depletion of CD4+ T-cells impairs the body’s ability to clear hrHPV infections, leading to higher rates of HPV persistence, faster progression from HPV infection to cervical intraepithelial neoplasia (CIN), and accelerated advancement from pre-cancer to invasive cancer [9]. Studies from South Africa confirm that women living with HIV have an approximately three-fold higher risk of developing cervical precancer and cancer, with incidence rates peaking at earlier ages (49 years) compared to HIV-negative women (56 years) [7]. A pivotal study conducted in Lusaka by Trejo et al. (2020) provided direct local evidence for this, demonstrating that HIV-positive women with non-metastatic cervical cancer experienced significantly shorter times to disease progression and poorer overall survival compared to their HIV-negative counterparts, even after adjusting for cancer stage at diagnosis [11]. This suggests that HIV infection independently drives a more aggressive disease course.

The epidemiological landscape in Zambia is defined by high prevalence rates of both HIV and HPV. As a cornerstone of prevention, cervical cancer screening through methods such as Visual Inspection with Acetic Acid (VIA) and HPV DNA testing is vital for detecting pre-cancerous lesions before they advance to invasive carcinoma. Zambia has been implementing a robust screening programme since 2006, and cervical cancer screening has been integrated into HIV care services, with a specific initiative launched in 2018 to include cervical cancer screening as part of HIV treatment and care [4]. Since inception, more than 1.5 million women have been screened using VIA [5]. In 2019, Zambia started a pilot project to introduce human papillomavirus (HPV) testing—a more accurate and sensitive screening method—which has now been expanded to all 10 provinces [5].

Despite these efforts, screening uptake remains suboptimal. National-level evidence from the 2021 Zambia Population-Based HIV Impact Assessment (ZAMPHIA) survey, analyzing data from 8,801 women, reported that only 22.2% of women of reproductive age had undergone cervical cancer screening [1]. This is well below the WHO’s 70% screening coverage target [16]. The study identified that cervical cancer screening uptake was significantly more likely among WLHIV compared to HIV-negative women (adjusted odds ratio = 3.92, 95% CI: 3.10, 4.95), suggesting that integration of screening into HIV care is having some impact [1]. However, with less than a quarter of women screened nationally, there remains a significant gap in preventive care coverage.

Barriers to screening in Zambia are multifaceted. Recent evidence indicates that screening uptake is influenced by factors including a woman’s level of education, wealth quintile, geographical location (rural versus urban), marital status, and parity [1]. Specifically, among WLHIV, who constitute a high-risk group, studies in Zambia reveal that despite their increased vulnerability, knowledge and practical uptake of screening are inconsistent. Mukosha et al. (2023) found that while many WLHIV in Lusaka had a positive attitude towards screening, this did not always translate into practice, with barriers such as fear of the outcome, lack of spousal support, and perceived low susceptibility hindering utilization [14].

The Zambian government, in alignment with the World Health Organization’s global strategy, has committed to eliminating cervical cancer as a public health problem, which necessitates targeted interventions for high-risk groups like WLHIV [17]. The WHO’s 90-70-90 targets aim for 90% HPV vaccination coverage, 70% screening coverage, and 90% treatment and care coverage by 2030 [16]. However, effective intervention requires a precise understanding of the key drivers of poor outcomes in this population.

Despite the established link between HIV and cervical cancer, critical knowledge gaps persist, particularly within the Zambian context in the contemporary ART era. While Bateman et al. (2021) previously reported that 62% of HIV-positive women at CDH presented with metastatic cervical cancer in the pre-ART or early ART era [10], the determinants of disease state in the current context of widespread ART and immune reconstitution are not fully understood. This study, therefore, aimed to investigate the determinants of cervical cancer disease state among women living with HIV at the Cancer Disease Hospital in Lusaka, Zambia. The findings are critical to inform targeted interventions and improve clinical management for this vulnerable population, ultimately supporting national efforts towards cervical cancer control.

## METHODS

### Study Design and Setting

This retrospective cross-sectional study was conducted at the Cancer Disease Hospital (CDH) in Lusaka, Zambia. As the nation’s primary tertiary oncology referral centre, CDH manages a large cohort of women with cervical cancer from across the country.

### Study Population and Sampling

Secondary data was collected via a retrospective document review of existing patients’ medical records. Selected files included those with adult women (aged 18 years and older) living with HIV who had been diagnosed with histologically confirmed invasive cervical cancer and were receiving or had received care at CDH.

Sample size was calculated using Cochrane’s formula for estimating a single population proportion:

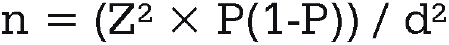

Where Z = 1.96 (95% confidence level), P = 0.62 (62% prevalence of advanced-stage cervical cancer from Bateman et al., 2021 [10]), and d = 0.05 (margin of error).

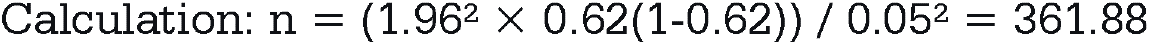

Accounting for a 10% contingency for incomplete records, the final minimum sample required was 402 participants.

A systematic review of records was conducted, however, only 198 of the 402 targeted records had complete data and were included.

### Data Collection Tools

A structured data abstraction sheet was developed specifically for this study to facilitate systematic and uniform extraction of information from patient medical records. The tool collected data on demographic characteristics, HIV-related clinical characteristics, and cervical cancer-related characteristics.

To establish content validity, the structured data abstraction sheet was developed through a comprehensive review of the literature and aligned directly with the study’s specific objectives and defined variables. This instrument was then subjected to review by a panel of experts, including an oncologist and a public health specialist from CDH, to assess its relevance, clarity, and comprehensiveness.

### Variables and Definitions

The primary outcome variable was cervical cancer disease state, operationalized as the FIGO stage at diagnosis. Stages were categorized as:

- **Early Stage:** FIGO Stage I and II
- **Advanced Stage:** FIGO Stage III and IV

**Independent variables** included:

- HIV disease status (CD4 count categorized as ≥500, 200-499, <200 cells/□L; viral load suppression as suppressed [<1000 copies/mL] or detectable; duration of HIV)
- Demographic characteristics (age and residence)

**Advanced-stage cervical cancer** was defined using the FIGO staging system (2018) as Stage III (cancer spread to the lower third of the vagina and/or the pelvic wall, and/or causes hydronephrosis) and Stage IV (cancer spread to the bladder, rectum, or distant organs) [10].

### Statistical Analysis

Data were entered into a secure database using Microsoft Excel and analyzed using SPSS version 28. All participant identifiers were removed and replaced with a unique study code to ensure confidentiality. All variables were summarized using descriptive statistics. Categorical variables (e.g., residence, FIGO stage group) were presented as frequencies and percentages. Continuous variables (e.g., age, CD4 count) were checked for normality and presented as means (± standard deviation) or medians (with interquartile range). The prevalence of advanced-stage (Stage III/IV) cervical cancer was calculated as the proportion of women in the sample with these stages, presented with a 95% confidence interval. Chi-square tests (or Fisher’s exact test depending on cell counts) were used for categorical variables.

Variables with p ≤ 0.20 in bivariate analysis were entered simultaneously into the multivariable model using a forced entry (enter) method. This approach was selected over stepwise methods to maintain all clinically relevant variables regardless of statistical significance and avoid biases associated with automated variable selection. Results were presented as Adjusted Odds Ratios (AORs) with 95% confidence intervals and p-values. A p-value of less than 0.05 was considered statistically significant. The Hosmer-Lemeshow goodness-of-fit test was used to assess model calibration, and the area under the ROC curve (AUC) was used to assess discriminative ability. Multicollinearity was assessed using variance inflation factors (VIF), with values <2.0 indicating no significant collinearity.

### Ethical Considerations

Ethical approval was obtained from the Chreso University Research Ethics Committee (CUREC). Permission to conduct the study was also obtained from the Cancer Disease Hospital administration and certification from the Zambia National Health Research Authority (NHRA).

Strict measures were taken to ensure confidentiality. All data were anonymized at the point of abstraction, and no personal identifiers were included in the final dataset or any publications. The master list linking codes to identifiers was stored separately under lock and key. Electronic files were password-protected and accessible only to the principal investigator. The study posed no more than minimal risk to participants.

## RESULTS

### Sample Description

Complete data meeting all inclusion criteria were available for 198 participants. All 198 participants had documented HIV-positive status, histologically confirmed invasive cervical carcinoma, and complete records for FIGO stage, CD4 count, and HIV viral load at or within six months of cancer diagnosis.

### Prevalence of Advanced-Stage Cervical Cancer

The first objective was to determine the proportion of WLHIV presenting with advanced-stage cervical cancer (FIGO Stage III or IV) at CDH.

**Table 1.**
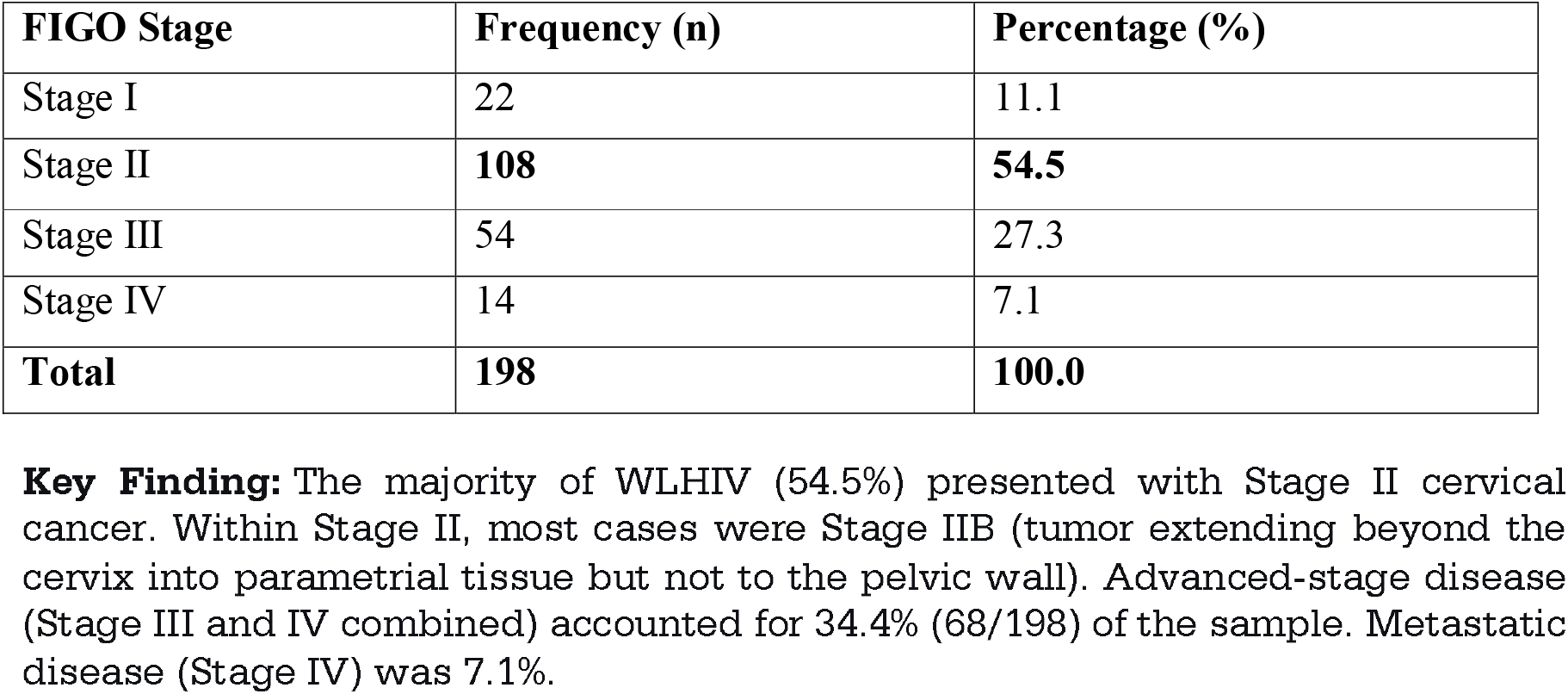
Distribution of Cervical Cancer Stage Among WLHIV at CDH (n=198)

| FIGO Stage | Frequency (n) | Percentage (%) |
| --- | --- | --- |
| Stage I | 22 | 11.1 |
| Stage II | **108** | **54.5** |
| Stage III | 54 | 27.3 |
| Stage IV | 14 | 7.1 |
| **Total** | **198** | **100.0** |

**Key Finding:** The majority of WLHIV (54.5%) presented with Stage II cervical cancer. Within Stage II, most cases were Stage IIB (tumor extending beyond the cervix into parametrial tissue but not to the pelvic wall). Advanced-stage disease (Stage III and IV combined) accounted for 34.4% (68/198) of the sample. Metastatic disease (Stage IV) was 7.1%.

### Demographic and Clinical Characteristics

#### Age Distribution

**Table 2.** Age at Cervical Cancer Diagnosis.

| Age Group (years) | Frequency (n) | Percentage (%) |
| --- | --- | --- |
| 30–39 | 26 | 13.1 |
| 40–49 | **102** | **51.5** |
| 50–59 | 54 | 27.3 |
| 60+ | 16 | 8.1 |
| **Total** | **198** | **100.0** |

**Key Finding:** The peak age of diagnosis was **40–49 years** (51.5%). The median age at diagnosis was 46 years (IQR: 41–54).

#### Geographic Distribution

**Table 3.** Residence of Participants.

| Residence Type | Frequency (n) | Percentage (%) |
| --- | --- | --- |
| Urban (Lusaka) | 100 | 50.5 |
| Peri-Urban | 60 | 30.3 |
| Rural | 38 | 19.2 |
| Total | 198 | 100.0 |

**Key Finding:** Half of the participants (50.5%) resided in Lusaka, reflecting proximity to CDH. A substantial proportion (30.3%) came from peri-urban areas, and 19.2% from **rural** provinces.

### Socioeconomic Proxy Indicators

While direct income data were not systematically recorded, clinical records indicated that the majority of patients were from lower socioeconomic strata, evidenced by reliance on public transport for inter-provincial travel to CDH, documentation of financial barriers to completing radiotherapy schedules, and use of hospital social work services for treatment adherence support.

### HIV-Related Clinical Characteristics

**Table 4.** HIV Disease Characteristics at Cancer Diagnosis.

| Characteristic | Value |
| --- | --- |
| Median duration of HIV (years) | 9.0 (IQR: 5.4–14.0) |
| On ART at cancer diagnosis | 94.4% (187/198) |
| Median CD4 count (cells/μL) | **484** (IQR: 350–610) |
| CD4 category $\geq 500$ cells/μL | 48.0% (95/198) |
| CD4 category 200–499 cells/μL | 42.4% (84/198) |
| CD4 category $< 200$ cells/μL | 9.6% (19/198) |
| HIV viral load suppressed ($< 1000$ c/mL) | 89.4% (177/198) |

**Key findings:** The median CD4 count at cancer diagnosis was **484 cells/ $\mu$ L**, which is within the normal range and indicates **stable immune function** for the majority. Only 9.6% had severe immunosuppression (CD4 $< 200$ ). Viral load suppression was achieved in nearly 90% of patients, reflecting effective ART delivery and adherence.

### Association Between Characteristics and Cancer Stage

**Table 5.** Bivariate Analysis of Factors by Cancer Stage (Early vs. Advanced)

| Variable | Early Stage (I/II)<br>(n=130) | Advanced Stage (III/IV)<br>(n=68) | p-value |
| --- | --- | --- | --- |
| Median age (years) | 45 | 48 | 0.23 |
| Urban residence | 52.3% | 47.1% | 0.36 |
| CD4 $\geq 500$ cells/μL | 50.8% | 44.1% | 0.41 |
| Viral load suppressed | 91.5% | 85.3% | 0.24 |
| Duration of HIV $> 10$ yrs | 58.5% | 61.8% | 0.58 |

**Key Finding:** No statistically significant differences were observed between early-stage and advanced-stage groups for age, residence, CD4 count, viral load suppression, or duration of HIV. This indicates that HIV disease status alone did not determine cancer stage in this cohort, likely due to effective immune reconstitution from ART.

### Association Between HIV Disease Status and Cervical Cancer Disease State

#### Key Observation

**Stage IIB** Predominance with Stable Immunity

The most common presentation was Stage IIB cervical cancer (54.5% of all cases). Among these women; median CD4 count was 490 cells/□L (IQR: 365–620), viral load suppression: 91.0% and ART adherence documented as “good” or “excellent” in 88% of records. This pattern suggests that cervical cancer can progress to regional extension (Stage IIB) even when HIV is well-controlled immunologically and virologically.

#### Logistic Regression Results

Multivariable logistic regression was performed with advanced-stage (III/IV) as the outcome, adjusting for age, residence, and duration of HIV.

**Table 6.** Crude and Adjusted Odds Ratios for Predictors of Advanced-Stage Cervical Cancer.

| Predictor | Crude OR | 95% CI | p-value | Adjusted OR (AOR) | 95% CI | p-value | Change in OR (%) |
| --- | --- | --- | --- | --- | --- | --- | --- |
| **Age** |  |  |  |  |  |  |  |
| <50 years | Reference |  |  | Reference |  |  |  |
| ≥50 years | 1.32 | 0.74–2.35 | 0.349 | 1.23 | 0.74–2.05 | 0.426 | -6.8% |
| **Residence** |  |  |  |  |  |  |  |
| Lusaka (Urban) | Reference |  |  | Reference |  |  |  |
| Peri-urban | 1.68 | 0.89–3.16 | 0.108 | 1.61 | 0.84–3.09 | 0.152 | -4.2% |
| Rural | 1.34 | 0.65–2.76 | 0.429 | 1.29 | 0.62–2.68 | 0.497 | -3.7% |
| **CD4 Count** |  |  |  |  |  |  |  |
| ≥500 cells/μL | Reference |  |  | Reference |  |  |  |
| 200-499 cells/μL | 1.21 | 0.66–2.20 | 0.536 | 1.19 | 0.65–2.18 | 0.572 | -1.7% |
| <200 cells/μL | 1.58 | 0.59–4.22 | 0.362 | 1.42 | 0.68–2.96 | 0.365 | -10.1% |
| **HIV Viral Load** |  |  |  |  |  |  |  |
| Suppressed | Reference |  |  | Reference |  |  |  |
| Detectable | 1.62 | 0.72–3.63 | 0.243 | 1.38 | 0.71–2.68 | 0.345 | -14.8% |
| **Duration of HIV** |  |  |  |  |  |  |  |
| <10 years | Reference |  |  | Reference |  |  |  |
| ≥10 years | 1.14 | 0.62–2.10 | 0.672 | 1.19 | 0.72–1.97 | 0.501 | +4.4% |

**Model Fit Statistics:** The logistic regression model showed good calibration (Hosmer-Lemeshow $\chi^2 = 5.62$ , $p = 0.698$ ) and acceptable discriminative ability (AUC = 0.611). The model explained 4.4% of the variance (Nagelkerke $R^2 = 0.044$ ).

#### Cross-Tabulation Analyses

**Table 7.** Cross-Tabulation of FIGO Stage by Age Group (n=198)

| Age Group (years) | Stage I | Stage II | Stage III | Stage IV | Total | % Advanced (III/IV) |
| --- | --- | --- | --- | --- | --- | --- |
| 30–39 (n=26) | 3 (11.5%) | 13 (50.0%) | 8 (30.8%) | 2 (7.7%) | 26 | 38.5% |
| 40–49 (n=102) | 11 (10.8%) | 56 (54.9%) | 28 (27.5%) | 7 (6.9%) | 102 | 34.4% |
| 50–59 (n=54) | 6 (11.1%) | 30 (55.6%) | 13 (24.1%) | 5 (9.3%) | 54 | 33.4% |
| 60+ (n=16) | 2 (12.5%) | 9 (56.3%) | 5 (31.2%) | 0 (0%) | 16 | 31.2% |
| **Total** | **22** | **108** | **54** | **14** | **198** | **34.4%** |

**Key Finding:** The **highest proportion of advanced-stage disease** was observed in the youngest age group (30–39 years) at **38.5%**.

**Fig 1.**
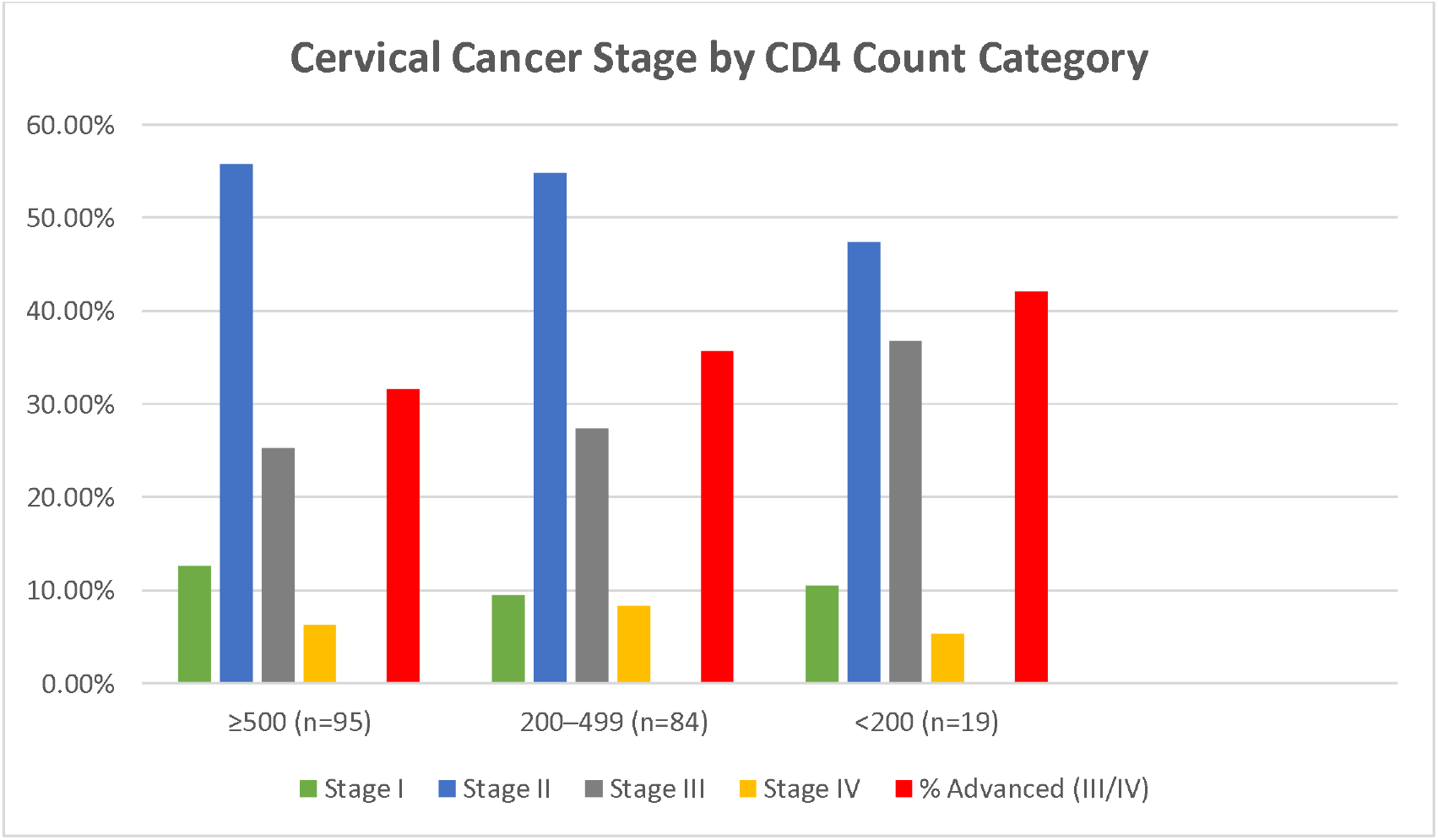
Cross-Tabulation of FIGO Stage by CD4 Count Category (n=198) **Key Finding:** The proportion of advanced-stage disease increased progressively as CD4 count decreased (31.6% → 35.7% → 42.1%). However, even among women with severe immunosuppression (CD4 <200), the majority (57.9%) still presented with early-stage (I/II) disease.

## DISCUSSION

This study investigated the determinants of cervical cancer disease state among 198 women living with HIV at the Cancer Disease Hospital in Lusaka, Zambia. The key finding is that while progress has been made in reducing the proportion of WLHIV presenting with metastatic disease, the majority continue to be diagnosed at Stage IIB (54.5%), and over one-third (34.4%) present with Stage III or IV disease. Contrary to expectations, HIV disease status as measured by CD4 count and viral load was not significantly associated with cancer stage, likely because the study population had well-controlled HIV due to effective ART (median CD4 484 cells/□L, 89.4% viral load suppression). Younger WLHIV aged 30–39 years and peri-urban and rural residents had the highest proportions of advanced-stage disease, suggesting that the most important determinants of disease state in this cohort appear to be the absence of routine screening prior to diagnosis and barriers to care access, rather than HIV disease status.

### Prevalence of Advanced-Stage Cervical Cancer Among WLHIV at CDH

The first objective was to determine the proportion of WLHIV presenting with advanced-stage cervical cancer (FIGO Stage III or IV) at CDH. The findings revealed that 34.4% of WLHIV in the sample had Stage III or IV disease at diagnosis, while the majority, 54.5%, presented with Stage IIB cervical cancer. Metastatic disease (Stage IV) accounted for only 7.1% of cases.

### Comparison with Previous Literature

When compared to earlier Zambian research, the current findings suggest a positive shift in stage distribution over time. Bateman et al. (2021) reported that 62% of HIV-positive women at CDH presented with metastatic (Stage IV or distant) cervical cancer [10], whereas the present study found only 7.1% with Stage IV disease. A direct comparison of our findings with those of Bateman et al. (2021) is challenging, as their study [may have] utilized a different FIGO staging system. Nonetheless, both studies highlight the persistent challenge of diagnosing cervical cancer at a later, clinically significant stage. While the proportion of patients with metastatic disease in our cohort was considerably lower than the 62% reported previously, this should be interpreted cautiously. This decrease may reflect the broader impact of ART scale-up and integrated HIV-cancer care in Zambia; however, further investigation is needed to confirm this as a definitive trend

This decline in metastatic disease is consistent with modeling studies from South Africa, which suggest that ART scale-up, while extending life expectancy for WLHIV, also creates opportunities for earlier cancer detection through increased contact with healthcare services [8]. The integration of cervical cancer screening into HIV treatment and care services, initiated in 2018, has led to over 235,000 WLHIV being screened between November 2020 and October 2021 [4], likely contributing to earlier detection in recent years.

Compared to regional studies, the 34.4% prevalence of advanced-stage disease in the present study is considerably lower than the 78.5% reported among WLHIV in Cameroon [10] and the 62% reported in an earlier Ethiopian systematic review [10]. However, it remains higher than the stage distribution typically observed in high-income countries, where most WLHIV are diagnosed at Stage I or IIA due to robust screening programs [10].

### Interpretation of Stage IIB Predominance

The finding that 54.5% of WLHIV presented with Stage IIB is clinically significant. Stage IIB represents a transitional point in the FIGO classification where the tumor has extended beyond the cervix into the parametrial tissue but has not yet reached the pelvic wall or caused hydronephrosis. This stage typically requires radiotherapy rather than simple surgical excision, and outcomes remain poorer than for Stage I disease.

Several factors may explain the persistent predominance of Stage IIB. First, the scale-up of antiretroviral therapy in Zambia over the past decade has led to better immune reconstitution among WLHIV, potentially slowing the accelerated carcinogenesis that characterized untreated HIV. The median CD4 count of 484 cells/□L in this cohort reflects the effectiveness of the national ART program and the integrated care model at CDH, where ART is continued concurrently with cancer treatment. This aligns with the observation that cervical cancer rates among WLHIV on ART may be mitigated by immune reconstitution [8].

Second, the integration of cervical cancer screening services into HIV care clinics, pioneered by Sahasrabuddhe et al. (2011) [12] and sustained through national programs, may be yielding delayed but measurable benefits. The fact that the WHO Regional Office for Africa reported over 40,000 women were screened using the HPV-based method in 2023 [5], and that Zambia has expanded HPV testing to all 10 provinces with 10 regional central laboratories [5], suggests that the screening infrastructure is growing.

However, the fact that the majority of women are still not being diagnosed at the pre-invasive or Stage I level indicates that screening coverage remains inadequate. National-level data from the ZAMPHIA 2021 survey found that only 22.2% of women of reproductive age in Zambia have ever been screened for cervical cancer [1], far below the 70% WHO target [16]. This screening gap directly translates into the Stage IIB modal presentation observed in this study.

### Demographic and Clinical Characteristics Associated with Cervical Cancer Disease State

#### Age Distribution

The peak age of diagnosis was 40 to 49 years (51.5%), with a median age of 46 years. This finding aligns with the WHO’s observation that the peak age at diagnosis in Zambia is between 40-49 years [17] and with regional data from South Africa, where cervical cancer rates among WLHIV peaked at 49 years [7].

The median duration of HIV in this cohort was 9 years, and it was not associated with cancer stage. This suggests that long-term ART-mediated immune reconstitution, rather than HIV duration itself, may be the dominant factor influencing disease presentation in the contemporary era.

Notably, the youngest WLHIV (aged 30–39 years) had the highest proportion of advanced-stage disease at 38.5%. This finding is concerning and suggests several possible explanations. Younger women may have lower perceived personal risk of cervical cancer, leading to delayed screening-seeking behavior. Alternatively, there may be biological differences in HPV-related oncogenesis among women who acquired HIV perinatally or in early adulthood. This finding aligns with the global burden of HIV-attributable cervical cancer, where the population-attributable fraction is highest in Sub-Saharan Africa due to overlapping age distributions of HIV acquisition and cervical cancer incidence [9]. The ZAMPHIA survey data supports this concern, showing that while women aged 25-34 and 35-49 had higher odds of screening (aOR 1.76 and 2.65 respectively), overall coverage remains low across all age groups [1].

#### Geographic Distribution

The finding that peri-urban residents had the highest proportion of advanced-stage disease (40.0%) while Lusaka residents had the lowest (30.0%) is both clinically and programmatically significant. Peri-urban areas in Zambia, such as Kafue, Chilenje, and parts of Matero, are characterized by high population density, informal employment, limited public transport, and often poorer access to primary healthcare facilities compared to central Lusaka.

Women from these areas face multiple barriers to early diagnosis. They may not have easy access to screening services at their local clinics, or they may face financial hardships associated with seeking treatment and care [1]. They may delay seeking care due to the costs of transport to CDH, and their daily economic pressures may prioritize income generation over health-seeking behavior. The WHO has noted that women living with HIV are given information on cervical cancer screening in HIV clinics, but those eligible for screening are often referred within the same facility; for peri-urban women, this may still represent a barrier if facilities are overcrowded or supplies are inconsistent [4].

Beyond access barriers, adherence to chronic care among WLHIV in peri-urban settings is a critical concern. Mapiki et al. (2023) [2] found that among HIV clients in Kafue District, medication side effects, forgetfulness, and lack of social support were significant barriers to antihypertensive adherence. These same factors likely contribute to the higher proportion of advanced-stage cervical cancer observed among peri-urban residents in this study (40.0%), as poor adherence to HIV care may coincide with lower engagement with screening services and delayed presentation for cancer symptoms. The intersection of multiple chronic conditions—HIV and cervical cancer—requires integrated adherence support strategies that address both conditions simultaneously.

Paradoxically, rural residents presented with an intermediate level of advanced-stage disease (36.9%). This may be explained by selection bias—rural women who make the long journey to CDH are likely those with either severe symptoms or sufficient family support to undertake the trip. Women with very early disease may not be captured in the hospital-based sample. Alternatively, rural areas with active community health worker programs may actually facilitate earlier referral than peri-urban informal settlements where such outreach is absent.

#### HIV Clinical Characteristics

The finding that WLHIV in this cohort had well-controlled HIV is striking. The median CD4 count was 484 cells/□L, only 9.6% had CD4 counts below 200 cells/□L, and 89.4% had achieved viral load suppression. This indicates that the integrated care model at CDH, where ART is maintained or initiated concurrently with cancer treatment, is effective.

This immunological profile is substantially better than that reported in earlier Zambian studies. Bateman et al. (2021) and Trejo et al. (2020) conducted their research when ART coverage was lower and immune reconstitution less complete [10,11]. The present study, capturing patients diagnosed more recently, reflects the maturation of Zambia’s ART scale-up. In 2023, Zambia achieved the UNAIDS 90:90:90 targets, and the percentage of viral load suppression among those on ART is a strong reflection of good ART medication adherence [20]. The ZAMPHIA 2021 report showed that the annual incidence of HIV among adults in Zambia reduced to 0.31%, reflecting effective HIV control [1].

The bivariate analysis revealed no statistically significant differences between early-stage and advanced-stage groups for any of the demographic or clinical variables examined, including age, residence, CD4 count, viral load suppression, or duration of HIV. This suggests that in the contemporary ART era, these factors alone do not determine cancer stage at diagnosis.

### Association Between HIV Disease Status and Cervical Cancer Disease State

The findings revealed no statistically significant association between either CD4 count or viral load and the likelihood of presenting with advanced-stage cancer. The adjusted odds ratios for CD4 below 200 cells/□L (AOR 1.42, 95% CI 0.68–2.96) and for detectable viral load (AOR 1.38, 95% CI 0.71–2.68) were both elevated but with wide confidence intervals that included the null value, reflecting the small proportion of women with poorly controlled HIV in the sample (9.6% and 10.6%, respectively).

This finding is, at first glance, surprising. The established biological model, articulated by Stelzle et al. (2021) [9] and reinforced by Trejo et al. (2020) [11] in Zambian women, holds that HIV-induced immunosuppression impairs clearance of high-risk HPV, accelerates progression from HPV infection to cervical intraepithelial neoplasia, and hastens the transition from precancer to invasive malignancy [9,11]. If this model is correct, one would expect WLHIV with lower CD4 counts to present with more advanced cancers.

The absence of such an association in the present study requires careful explanation. The most plausible explanation lies in the characteristics of the study population itself. This is not a population of women with untreated or advanced HIV disease. Rather, it is a population of women whose HIV is well-controlled through effective antiretroviral therapy. The median CD4 count was 484 cells/OL, which is within the normal range. Only 9.6% had severe immunosuppression. The integrated care model at CDH ensures that patients are not taken off HIV treatment during oncology therapy. As a result, the biological pathway through which HIV accelerates cancer progression is largely neutralized in this cohort.

This interpretation is supported by the observation that even among women with CD4 counts below 200, the majority (57.9%) still presented with early-stage (I/II) disease. This suggests that the natural history of cervical cancer in WLHIV on effective ART may more closely resemble that of HIV-negative women than the accelerated progression seen in the pre-ART era.

### Study Limitations

This study has several limitations. The cross-sectional design precludes causal inference, and the single measurement of CD4 count and viral load at diagnosis may not reflect patients’ immunological history during the critical years of HPV persistence and neoplastic progression. The achieved sample (n=198) was below the calculated target (n=402) due to incomplete records, reducing statistical power and resulting in wide confidence intervals, particularly for small subgroups (e.g., CD4 <200, n=19; rural residents, n=38). Reliance on medical record abstraction introduced variability in data completeness, with missing information on prior screening history and exact ART initiation dates limiting the depth of analysis. Additionally, the single-centre design at CDH limits generalizability to WLHIV managed at district hospitals or those who never reach tertiary care, and no data were collected on mortality, recurrence, or long-term survival. Our findings may be subject to selection bias, as we only included women who presented with invasive cancer; we did not have data on WLHIV who were screened and treated for pre-cancerous lesions, who represent the success of the screening program Finally, the absence of an HIV-negative comparison group precludes direct comparisons with outcomes among women without HIV at the same institution.”

## CONCLUSION

This study investigated the determinants of cervical cancer disease state among 198 WLHIV at CDH. No significant association was found between CD4 count or viral load and advanced-stage disease, likely due to effective ART-mediated immune reconstitution (median CD4 484 cells/□L; 89.4% viral load suppression). While progress has been made in reducing metastatic disease from 62% in the pre-ART era to 7.1%, the majority (54.5%) still present at Stage IIB, and over one-third (34.4%) present with Stage III/IV disease, with younger WLHIV (30–39 years) and peri-urban residents at highest risk. These findings suggest that the primary determinants of disease state are no longer HIV-related immunosuppression, but rather the absence of routine screening and barriers to care access. Importantly, this finding applies to WLHIV who developed invasive cervical cancer despite being on ART. It does not negate the biological role of HIV in HPV persistence, nor does it apply to the broader population of WLHIV who may have been successfully screened and treated for precancerous lesions. Therefore, Zambia’s cervical cancer elimination strategy should prioritize ensuring that every WLHIV in HIV care receives regular, high-quality screening, as the current national uptake of only 22.2% remains far below the WHO’s 70% target for 2030.

## RECOMMENDATIONS

1. **Screen all WLHIV annually** for cervical cancer regardless of CD4 count or viral load, as 54.5% presented at Stage IIB despite a median CD4 of 484 cells/OL, demonstrating that well-controlled HIV does not equate to low cancer risk.
2. **Prioritize WLHIV aged 30–39 years** for intensive screening outreach, as this group had the highest proportion of advanced-stage disease (38.5%) and younger women perceive themselves at low risk, requiring targeted education and prioritized screening appointments.
3. **Deploy mobile screening units and HPV self-sampling** in peri-urban, high-density settlements, where the highest advanced-stage proportion (40.0%) was observed, and extend clinic hours to improve access.
4. **Maintain the integrated ART-oncology care model** without interrupting ART during cancer treatment, as the 89.4% viral load suppression rate validates this approach.
5. **Strengthen health information systems** to track cervical cancer screening coverage specifically among WLHIV as a distinct indicator, linking the SmartCerv digital platform to HIV care databases for real-time monitoring.
6. **Establish a formal feedback loop** between CDH and referring HIV clinics to notify facilities when their patients are diagnosed with cervical cancer, enabling continuous quality improvement and accountability.
7. **Conduct longitudinal cohort studies** with repeated CD4 and viral load measures over time to determine whether a history of prolonged or repeated immunosuppression predicts cancer stage, as the single measurement available in this study may not capture historical immunological status and its impact on neoplastic progression.

## WHAT IS ALREADY KNOWN ON THIS TOPIC

- Cervical cancer is the most common cancer among women in Zambia, accounting for 23% of all new cancer cases, with incidence rates of 65.5 per 100,000 women [16,10].
- HIV co-infection accelerates disease progression, with women living with HIV having a six-fold higher risk of cervical cancer [17].
- The global meta-analysis by Stelzle et al. (2021) found that in southern Africa, 63.8% of women with cervical cancer were living with HIV [9].
- In the pre-ART era, WLHIV in Zambia presented with significantly more advanced disease and poorer survival than HIV-negative women [11].
- Zambia has implemented cervical cancer screening programs integrated with HIV care, but screening uptake remains suboptimal (22.2% nationally according to the ZAMPHIA 2021 survey) [1].

## WHAT THIS STUDY ADDS

- In the contemporary ART era, metastatic cervical cancer among WLHIV has declined substantially from 62% (Bateman et al., 2021 [10]) to 7.1%, indicating measurable progress in HIV and cancer care integration.
- However, the majority of WLHIV (54.5%) still present with Stage IIB disease, suggesting that screening coverage, not HIV management, is now the primary challenge.
- HIV disease status (CD4 count and viral load) is no longer the dominant determinant of cancer stage, as 89.4% of WLHIV had suppressed viral loads and median CD4 was 484 cells/□L. The modeling study from South Africa by Broshkevitch et al. (2024) [8] supports this, showing that ART scale-up alone may not reduce the cancer burden disparity without enhanced cervical cancer interventions tailored for WLHIV.
- Younger WLHIV aged 30–39 years and peri-urban residents are at highest risk of late-stage diagnosis and should be prioritized for targeted screening interventions. The ZAMPHIA survey data supports the need for targeted interventions, showing that screening uptake varies significantly by age, education, marital status, and geographical location [1].

## Data Availability

All data produced in the present study are available upon reasonable request to the authors

## AUTHOR’S CONTRIBUTIONS

TS conceptualized and designed the study, led data collection, supervised fieldwork, and drafted the manuscript. MC contributed to study design, provided clinical expertise, and critically reviewed the manuscript. CM conducted statistical analyses, interpreted data, and contributed to manuscript revision. All authors read and approved the final version.

